# Thyroid hormone axis and anthropometric recovery of children/adolescents with overweight/obesity: a scoping review

**DOI:** 10.1101/2022.01.17.22269437

**Authors:** Carlos Ramos Urrea, Amanda Paula Pedroso, Fernanda Thomazini, Andreia Cristina Feitosa do Carmo, Mônica Marques Telles, Ana Lydia Sawaya, Maria do Carmo Pinho Franco, Eliane Beraldi Ribeiro

## Abstract

Thyroid hormones play multiple physiological effects essential for the maintenance of basal metabolic rate (BMR), adaptive thermogenesis, fat metabolism, and appetite. The links between obesity and the hormones of the thyroid axis, i.e., triiodothyronine (T3), thyroxine (T4), thyrotropin-releasing hormone (TRH), and thyrotropin (TSH), are still controversial, especially when considering children and adolescents. This population has high rates of overweight and obesity and several treatment approaches, including nutritional, psychological, and physical exercise interventions have been used. Understanding the importance of the hormones of the thyroid axis in the recovery from overweight and obesity may help directing measures to the maintenance of a healthy body composition. The present scoping review was carried out to analyze studies evaluating these hormonal levels throughout interventions directed at treating overweight and obesity in children and adolescents. The main purpose was to ascertain whether the hormones levels vary during weight loss. We selected for analysis 16 studies published between 1999 and 2019. Most of the studies showed that the changes in body composition parameters in response to the multidisciplinary interventions correlated positively with free T3 (fT3)/ total T3 (TT3)/TSH. With respect to free T4 (fT4)/ total T4 (TT4), the most common finding was of unchanged levels and hence, no significant association with weight loss. Importantly, the response to the intervention has even been found to not be affected by fT4 supplementation. Further studies are necessary to elucidate the relevance of the variations in hormone levels to the establishment of overweight/obesity and to the recovery from these conditions in children/adolescents.

## 1. Introduction

Globally, obesity is a well-recognized public health problem affecting both adults and children(1). In children/ adolescents, the prevalence of overweight/obesity is high (2) and associates with increased risk to develop diabetes and other co-morbidities (3).

The pathophysiology of overweight/obesity includes genetic, environmental, behavioral, metabolic, psychological factors, and hormonal factors. Thyroid hormones play multiple physiological effects essential for the maintenance of basal metabolic rate (BMR), adaptive thermogenesis, fat metabolism, and appetite (4). The participation of the levels of the hormones of the thyroid axis, i.e., triiodothyronine (T3), thyroxine (T4), thyrotropin-releasing hormone (TRH), and thyrotropin (TSH), has been studied with no conclusive results, especially when considering children and adolescents. They have indicated either that thyroid-hormones resistance is a causal factor of obesity or that elevated hormone levels may represent an adaptive response to obesity (5).

The treatment of children and adolescents with overweight or obesity is a very relevant issue, and several approaches, including nutritional, psychological, and physical exercise interventions have been used (6). Understanding the importance of the hormones of the thyroid axis in the recovery from overweight and obesity may help directing measures to the maintenance of a healthy body composition.

The present scoping review was carried out to analyze studies evaluating these hormonal levels throughout interventions directed at treating overweight and obesity in children and adolescents.

## 2. Materials and Methods

This scoping review was registered on the International Prospective Register of Systematic Reviews (PROSPERO, CRD42020203359) and performed with the recommendations of Preferred Reporting Items for Systematic Reviews and Meta-Analyses extension for Scoping Reviews (PRISMA-ScR).

### 2.1. Eligibility criteria

Original articles published in peer-reviewed journals, written in English, Portuguese, or Spanish, that had children and/or adolescents with overweight or obesity as participants and that performed some intervention for weight management, including, nutritional and/or psychological, and/or medical, and/or exercise.

The exclusion criteria were systematic reviews, studies in animals or adults, use of growth hormone or steroid hormones, diagnostic of thyroid kidney, heart, or neurological illness.

### 2.2. Literature search

The electronic search in the databases was carried out based on a search strategy aiming at locating both published and unpublished studies. Data collection and analysis were performed in September and October 2020. Electronic searches were conducted using the following databases: MEDLINE way Pubmed, Latin American and Caribbean Literature in Health Sciences (LILACS), Scopus (Elsevier) and Cochrane Library.

The following descriptors were extracted from the Health Science Descriptors database: obesity, overweight, obese, excess weight, weight gain, malnutrition, thyroid hormones, thyroid concentrations, thyroid stimulating hormone, triiodothyronine, thyroxine, thyroid gland, child, children, adolescents. The development of the search strategy followed the recommendations of the checklist Peer Review of Electronic Search Strategies (PRESS) (7).

### 2.3. Study selection and appraisal

The selection and analysis of the studies were carried out by two independent authors using the Rayyan tool. The first selection was based on the title and summary of the studies. Duplicates and articles whose full texts were not available were excluded. Conflicts were resolved by consensus. After selection according to the inclusion criteria, two reviewers independently analyzed the texts in full to identify the relevant outcomes.

#### Data extraction and synthesis of results

To characterize the findings, the following variables were considered: age, type of intervention, effect of the intervention on body composition, and hormone levels, both at baseline and after the intervention. The articles were grouped by type of comparisons (in-group or between obese and eutrophic). Two studies involving thyroxine supplementation were constituted a third category.

## 3. Results

A total of 1031 articles were screened, leading to 20 eligible articles, of which 4 were excluded due to absence of full text. Sixteen articles were thus included in the qualitative synthesis. Figure 1 describes the selection process and Table 1 shows the results of each selected article.

**Figure 1.**
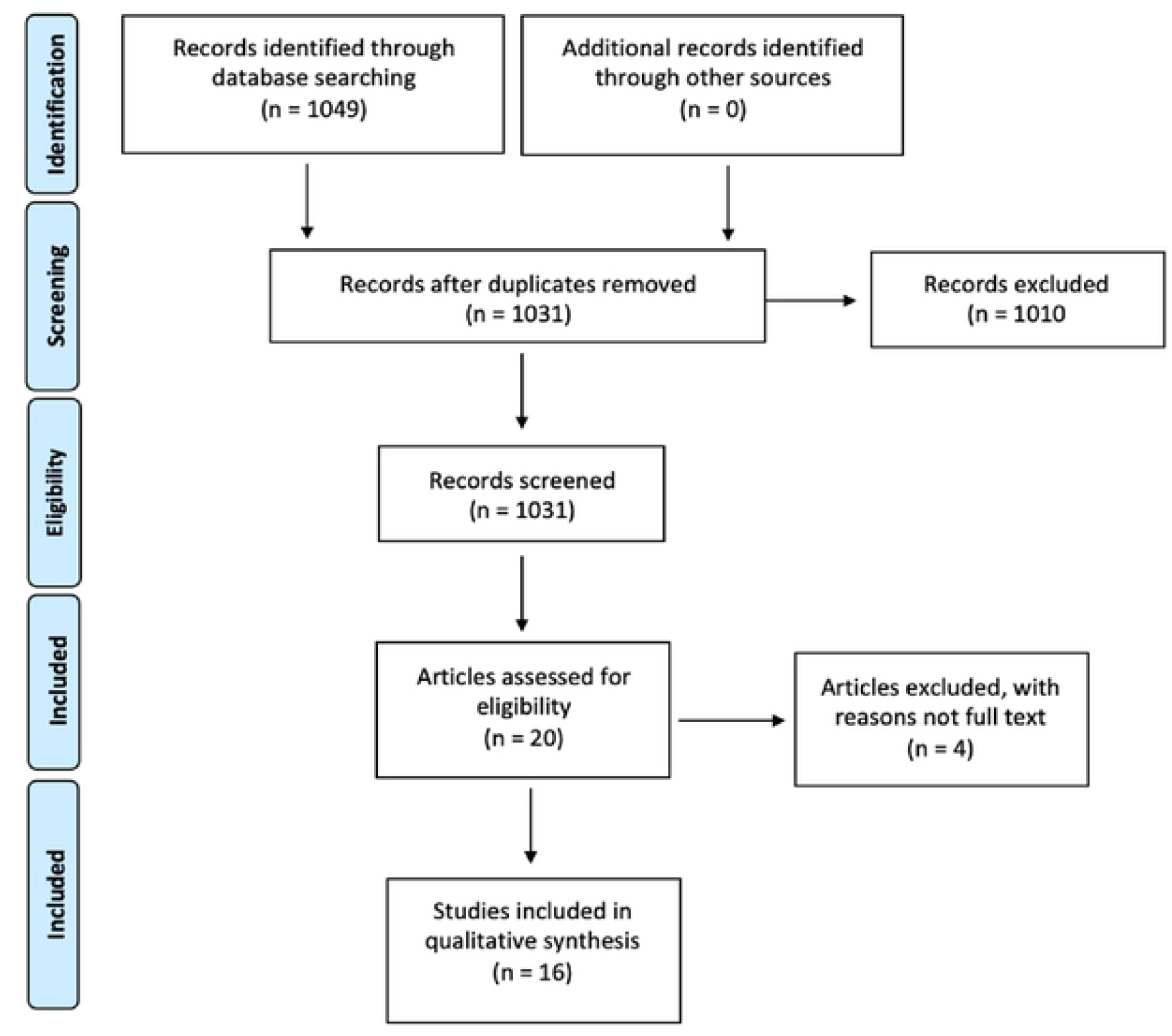
Flow diagram of the studies selection.

**Table 1.**
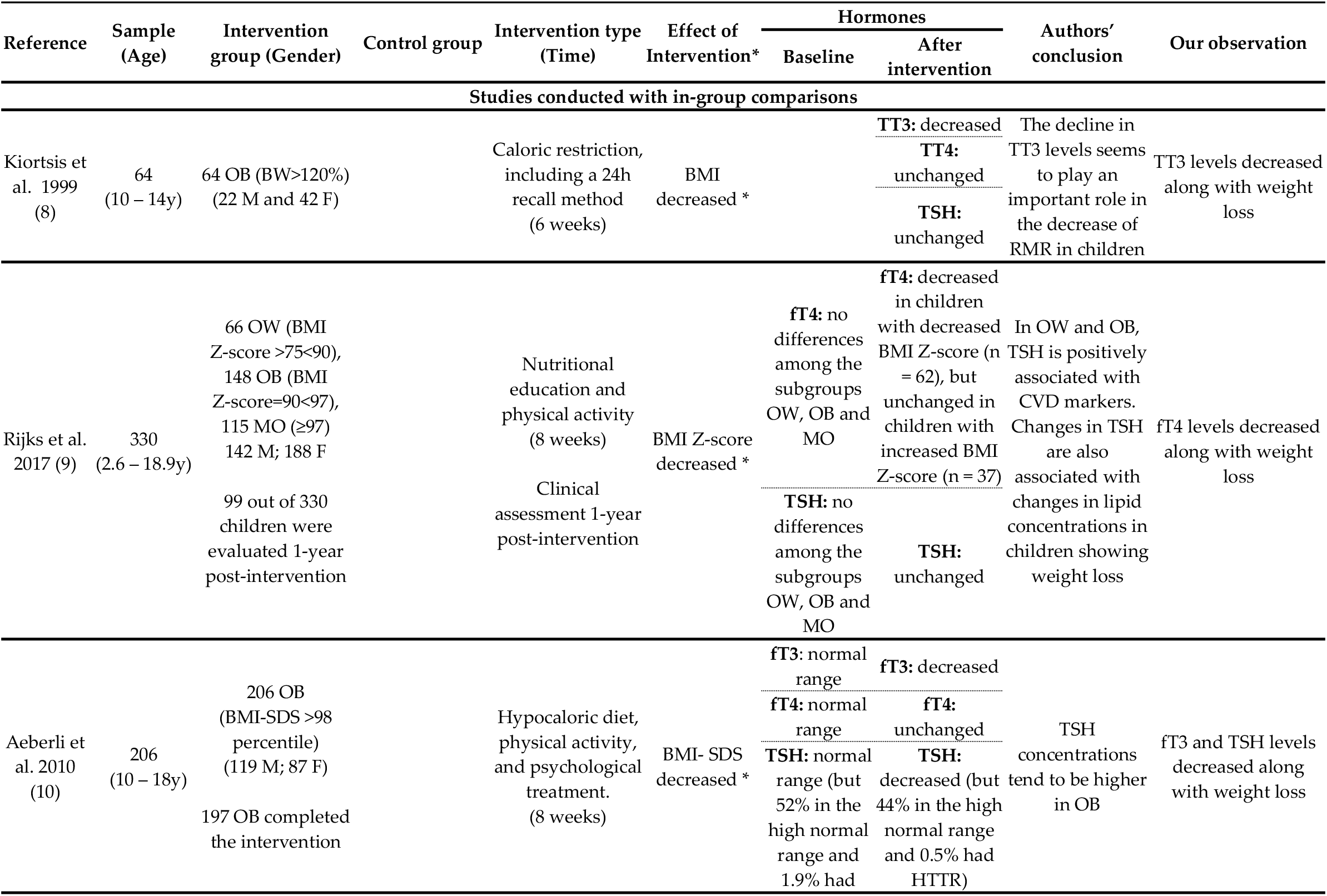
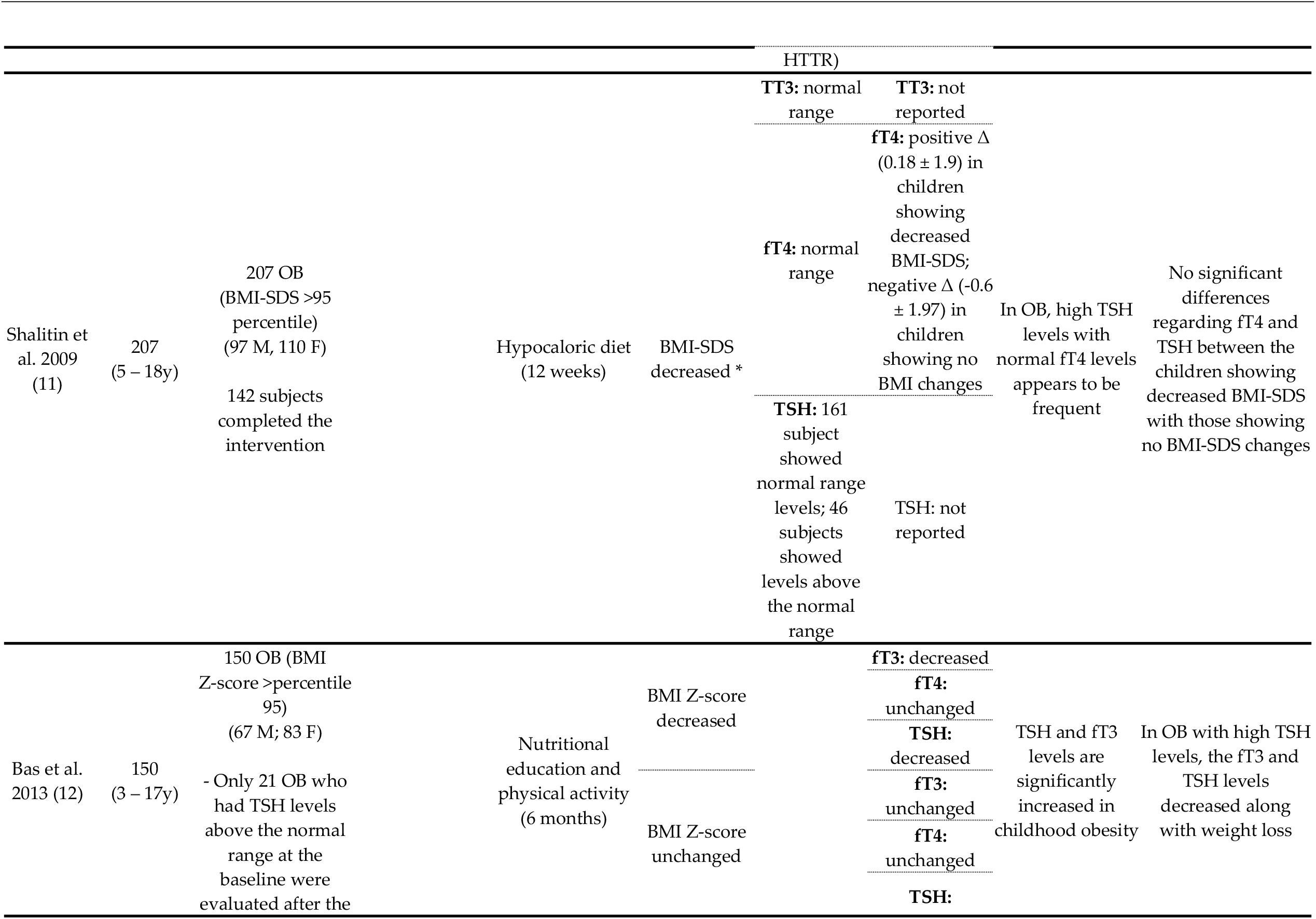
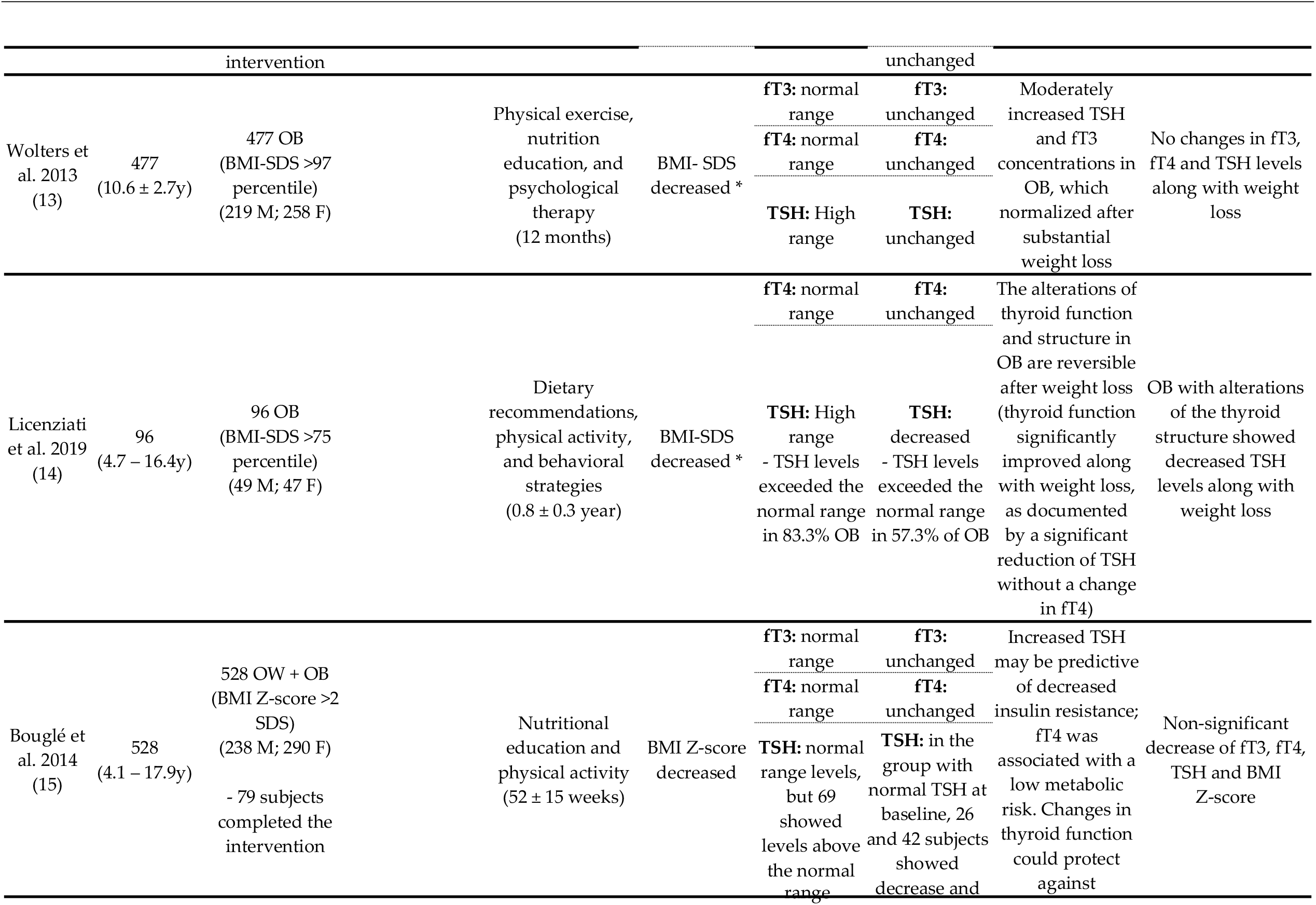
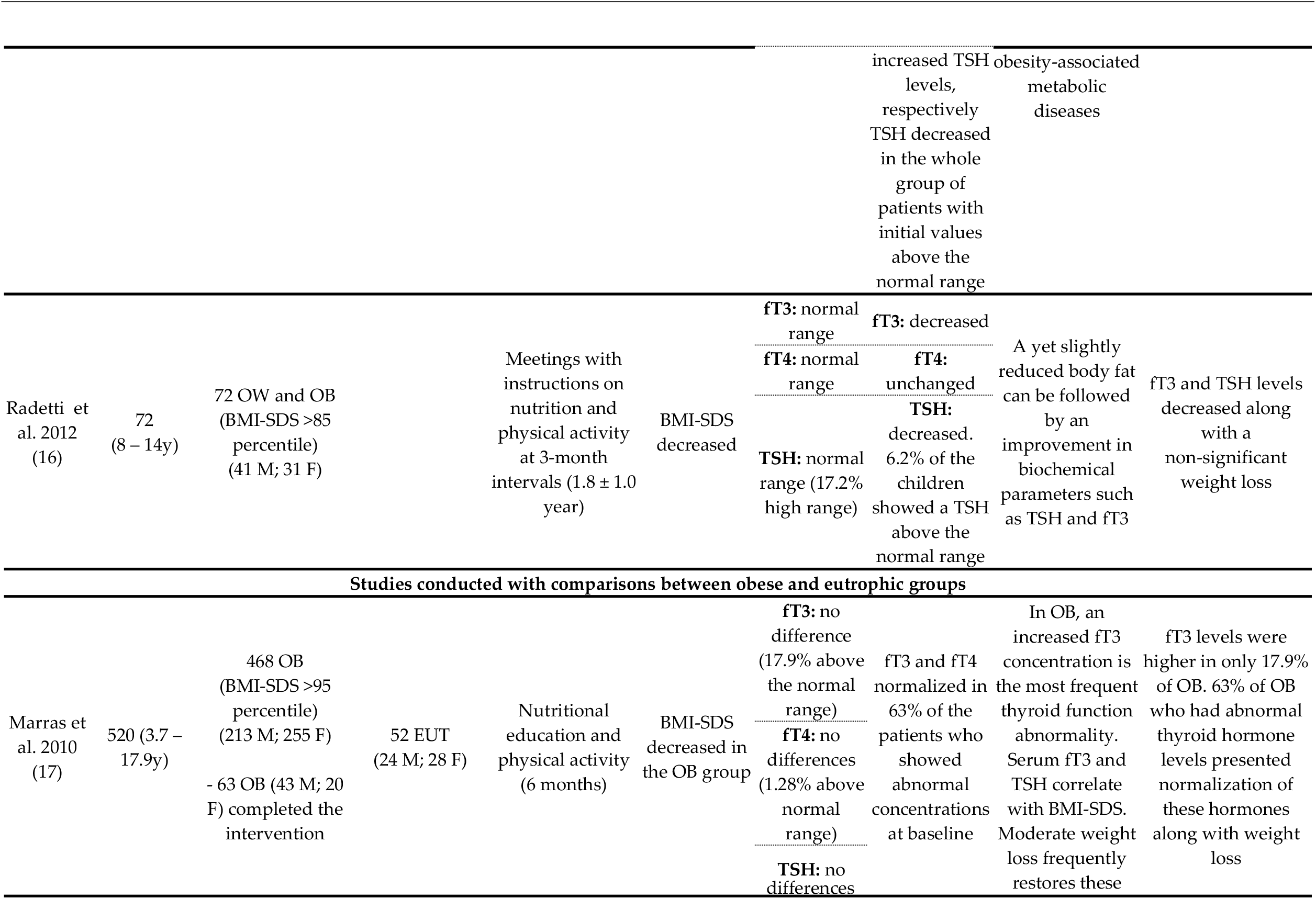
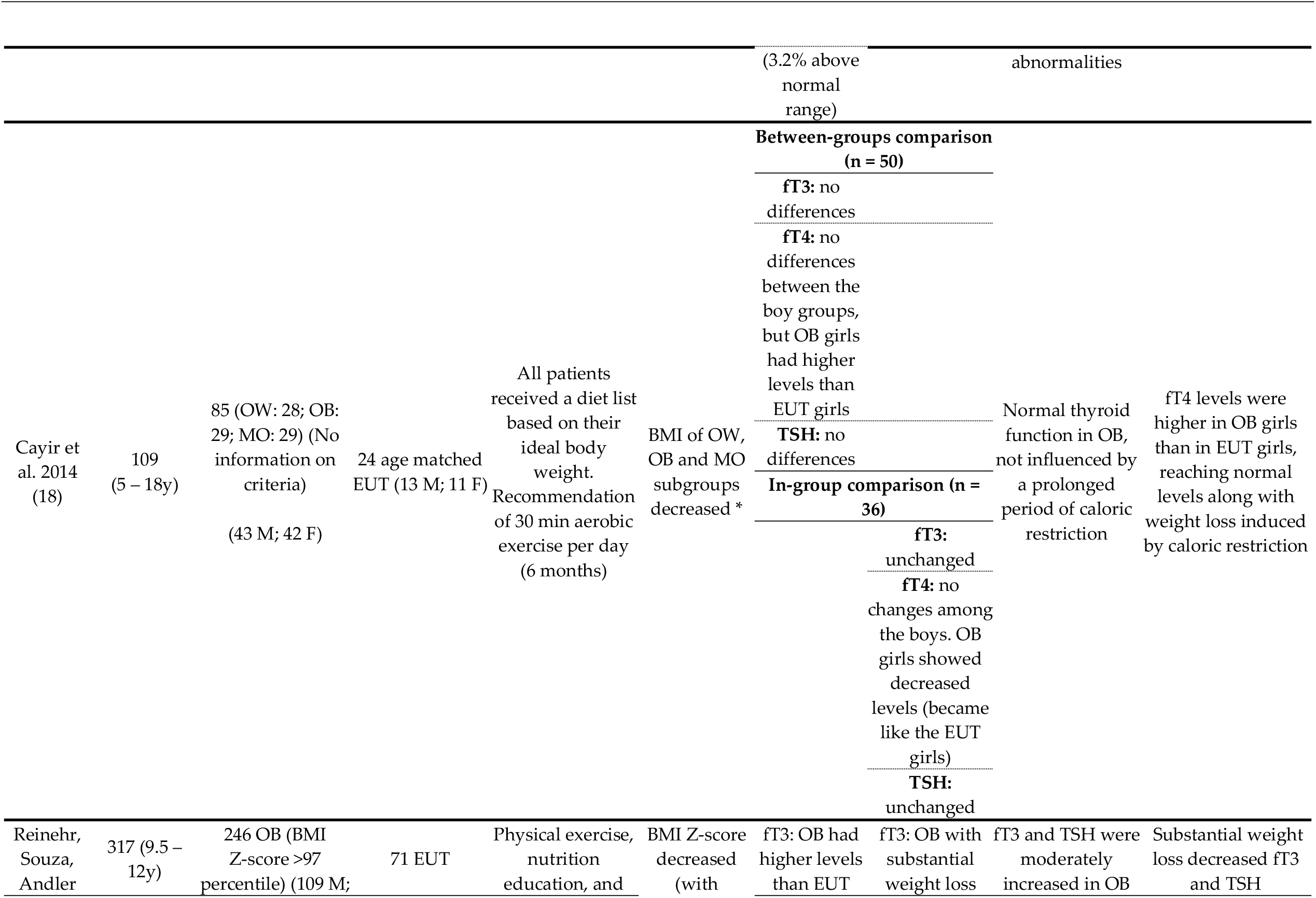
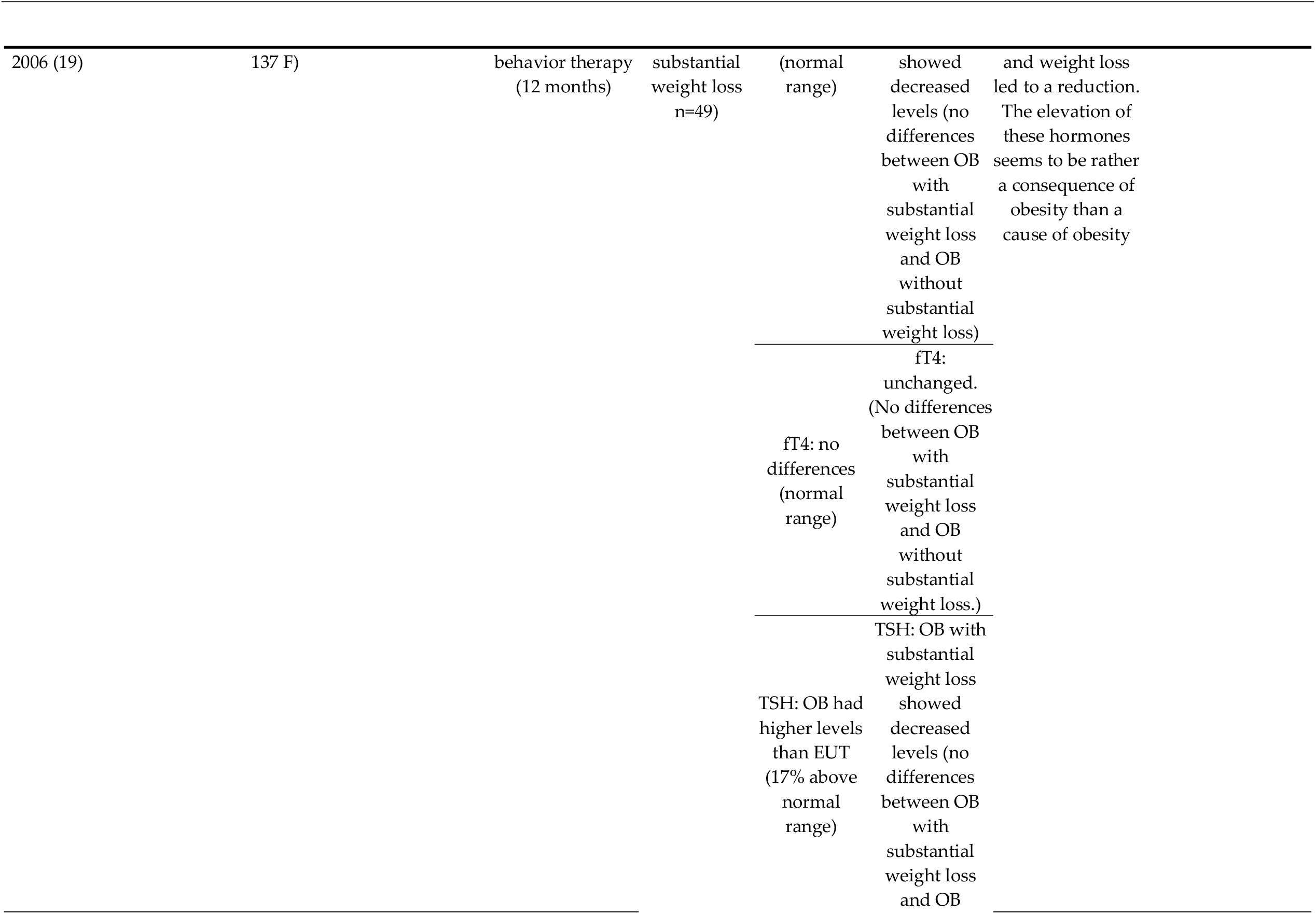
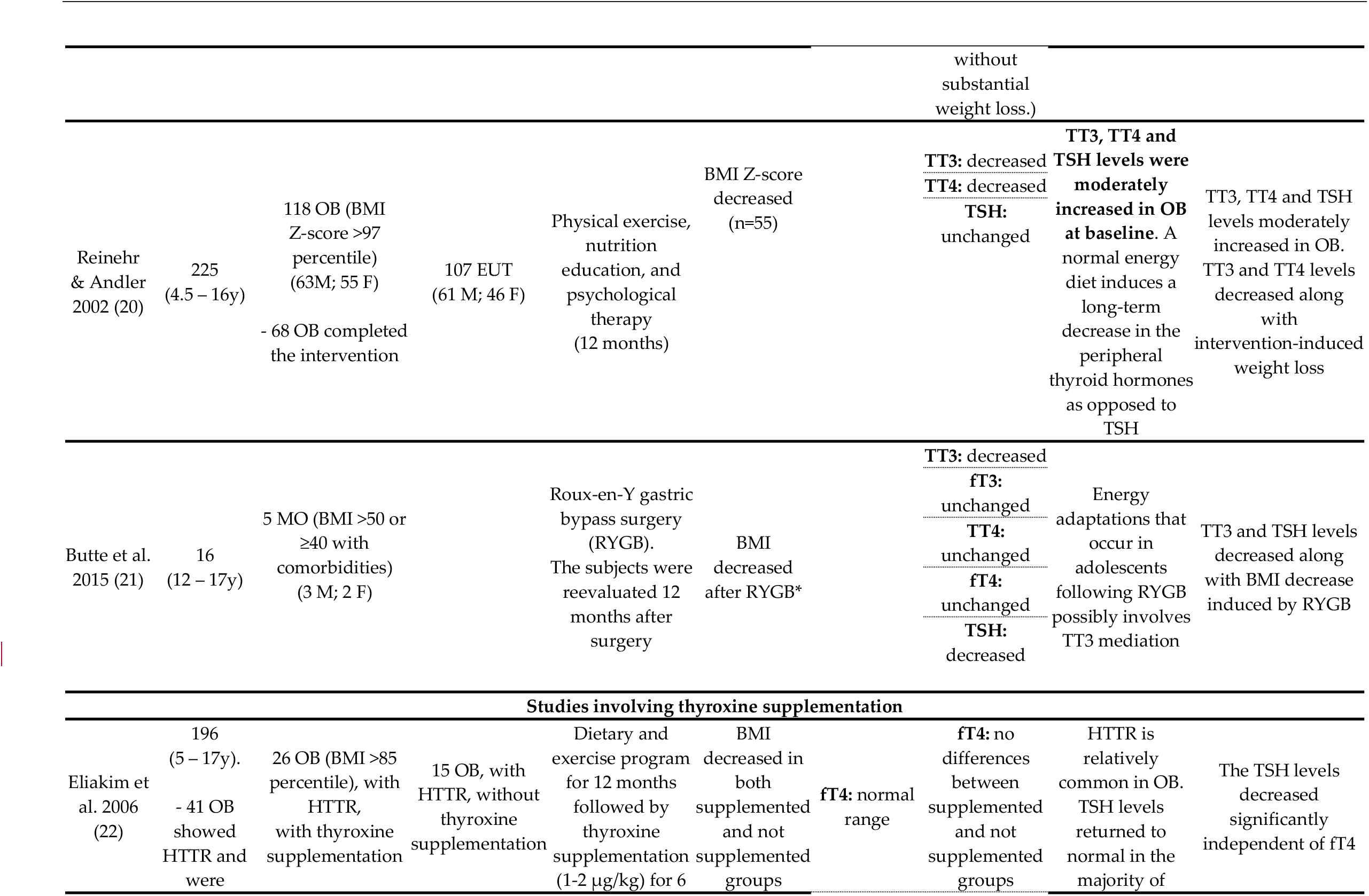
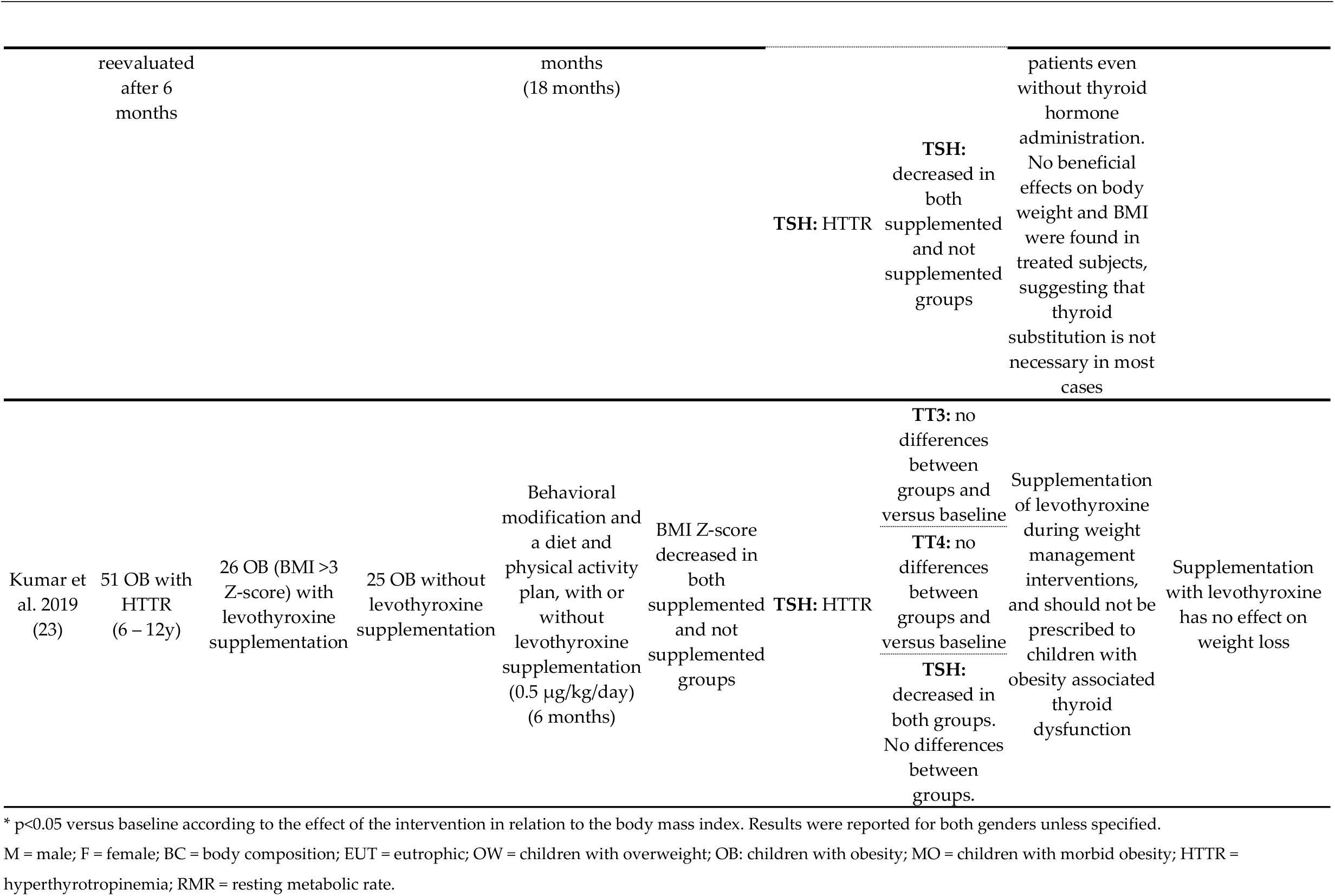
Summary of the characteristics and results of the 16 studies included in the analysis.

### 3.1. Description of the included studies

The literature search identified 1049 references (Figure 1) and 16 articles met all the inclusion criteria.

Table 1 summarizes the characteristics of the studies included in the analysis. There are studies performed with subjects from Italy (14,16,17), France (8,15), Israel (11,22), Turkey (12,18), Germany (13,19,20), the Netherlands (9), Swiss (10), United States (21), and India (23).

Twelve studies analyzed only obese children and adolescents (8,10–14,17,19–23) while the other 4 studies targeted both overweight and obese children and/or adolescents (9,15,16,18).

Table 2 describes the correlations between the thyroid hormonal axis and body composition parameters in the 9 studies that performed this calculation. At baseline, body weight was negatively correlated with fT3 (10), the percentage of fat mass was positively correlated with fT4 (10), and BMI was positively correlated with TSH (14,15,17) and fT3 (17). After intervention, BMI correlated positively with TSH, TT3, and TT4 (14,20,21). When considering the baseline-after intervention changes, the authors reported positive correlations of BMI and TT3 (8), and fT3 (10,12), and TSH (12). fT3 also correlated positively with body weight, fat mass, and percentage body fat (10). Negative correlations were seen between lean body mass and fT3 and percentage body fat and fT4 (10).

**Table 2.**
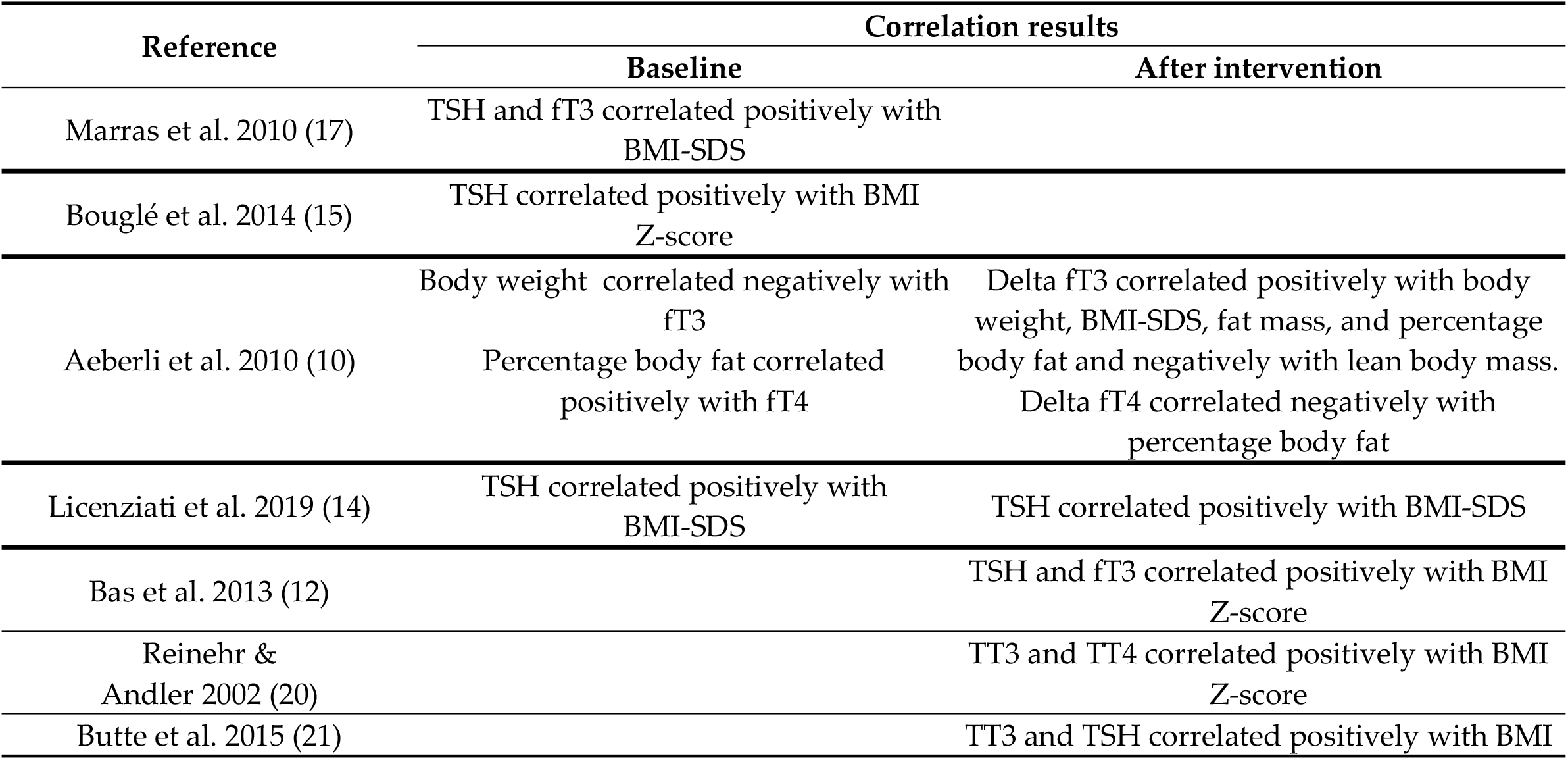
Correlations between anthropometric recovery and hormonal levels.

### 3.2. Interventions

The duration of the interventions varied from 6 weeks to 18 months. One study utilized only a nutritional intervention [8), 6 studies performed nutritional intervention plus exercise intervention (11,12,15–18). One study allied nutritional and exercise interventions to administration of thyroxine (22).

In 5 studies, a psychological intervention was added to the nutritional plus exercise intervention (10,13,14,19,20). One study performed the 3 interventions plus levothyroxine supplementation (23).

One study performed psychological and exercise interventions (9) and 1 study used only a surgical intervention (21).

### 3.3. Nutritional Interventions

Fourteen studies used nutritional interventions. Eight studies used a nutritional education (12–16,19,20,23), 5 studies used a calorie restriction approach, with energy levels ranging from 900 to 2000 kcal/d and with varied macronutrient combinations and 1 study used the Mediterranean diet (17).

### 3.4. Exercise Interventions

Thirteen studies utilized physical exercise interventions. Nine studies conducted supervised training sessions with no specific routine or duration, with varying types and intensities (9,12–16,19,20,23), 1 involved two daily group endurance exercise sessions to improve aerobic performance, with a typical session lasting 60–90 min (10), and 3 performed aerobic exercise 3–5 times/week for at least 45–60 min (17), 30 minutes per day (18), or 90 minutes per day (11).

### 3.5. Psychological intervention

This type of intervention was used in 6 studies, including individual psychological care of the child/adolescent (10,14) or of the child/adolescent and their family (9,13,19,20). In the 2 studies detailing the psychological intervention, it consisted of techniques focusing on increasing self-esteem, responsibilities, and problem-solving strategies (9) and relaxation techniques and breathing therapy (10).

## 4. Discussion

All the 16 studies included in this analysis, published between 1999 and 2019, reported anthropometric recovery of the overweight/obese children/adolescents in response to the interventions, which, as depicted in table 1, varied largely with respect to the type and duration.

Concerning the baseline levels of the thyroid hormones (fT3, fT4, TT3 or TT4), 6 studies did not report these data (8,9,12,20,21,23). Among the 10 studies in which this information was available, the majority (9 studies) reported no significant alterations, either in relation to the normality ranges (10,11,13–16,19,22) or in comparison to eutrophic individuals (17,18), although one of these latter studies reported higher levels of fT4 in obese than in eutrophic girls (18) and one study reported small percentages of subjects with levels of fT3 (17.9%) or fT4 (1.28%) above normal range. Only one study reported higher mean fT3 values in the obese than in the eutrophic subjects, although still in the normal range (19). These results show that the most common status of thyroid hormones falls into normal levels. TSH levels at baseline were not reported in 5 studies (8,9,12,20,21) while 2 studies selected only individuals with hyperthyrotropinemia (22,23). Among the remaining 9 studies, the mean levels were normal in 6 studies (10,11,15–19), although some of these studies found mean values in the high normal range and highlighted the presence of elevated levels in variable percentages of their cohorts, namely 1.9% (10), 28.6% (11), 13.1% (15), 17.2% (16), and 3.2% (17). Elevated baseline levels of TSH were reported in 3 studies (14,19,17). These data demonstrate that the most common status of TSH among the studies analyzed fell into normal levels, although the finding of values in the high normal range was frequent.

Four studies reported correlations between body measures and hormone levels at baseline. One study reported a positive association of fT3 with BMI-SDS (17) while another study found a positive association of fT3 and body weight and a negative association of fT4 and percentage body fat (10). In 3 studies, TSH correlated positively with BMI-SDS (14,17) and BMI Z-score (15).

We searched other studies reporting levels of the hormones of the thyroid axis in children/adolescents with overweight/obesity. In one study, no differences were found in the levels of fT4 and TSH between children/adolescents with excess weight and the eutrophic ones (13). Many studies showed that these levels felt into the normal range, although a common finding was that they were higher than those of eutrophic children/adolescents, concerning TT3 (24), TT4 (25), and TSH (24–28), fT3 and fT4 (5).

Similar findings have been found in adults, with respect to fT3 (29) and TT4 (29,30), i.e., levels in the normal range but higher than the eutrophic levels. There are also reports that the hormone levels were in the normal range but lower in obese than in eutrophic adults, concerning TSH (29–31), fT3 and fT4 (31).

Examining studies performing correlation analysis of hormone levels and body parameters in overweight/obese children/adolescents, we observed one study reporting no significant associations of fT3, fT4, and TSH levels with body composition parameters (32). In contrast, we found reports of a positive correlation between fT3 and BMI (5) and of a negative correlation of fT4 and BMI (25). There are also studies showing a positive correlation of TSH and body measures (5,33). These latter results agree with the findings of the studies analyzed in this scoping review.

These findings are like some reports in adults, finding positive associations between BMI and TT3 (34) and fT3 (29,31,34–38), and a negative correlation with fT4 (38). With respect to TSH, the studies found achieved a negative (30) or positive (38–41) association of BMI and TSH.

Concerning the response to the interventions, 10 of the studies included in this scoping review reported a decrease in at least one thyroid hormone measured (TT3, TT4, fT3, fT4) in relation to the respective baseline values (8–12,16,17,19– 21). Five studies did not find any type of significant change between the baseline and the post-intervention values (13– 15,18,23). One study did not report these data (22).

In relation to TSH, 9 studies reported a decrease after the intervention (10,12,14–16,19,21–23) and 5 studies showed no changes (8,9,13,18,20). It is important to point out that 2 of these studies included only subjects with hyperthyrotropinemia. In 2 studies, this information was not reported (11,17).

The relation of hormonal levels on anthropometric recovery was evaluated in 5 studies by a correlation analysis. Positive associations were found between delta of fT3 and body weight, BMI-SDS, fat mass, and percentage of fat mass (10), fT3 and BMI Z-score (12) and TT3 and BMI (21). TT4 correlated positively with BMI Z-score (20) but delta fT4 correlated negatively with percentage of fat mass (10). TSH correlated positively with BMI-SDS (14), BMI Z-score (12) and BMI (21). Studies performed in adults submitted to multidisciplinary interventions to treat obesity corroborate the above results, as they have found positive associations of fT3/TT3 with BMI and body weight (34,42,43) and of TSH with body weight (43–46). However, we found one study in which body and fat mass losses were not accompanied by in TSH levels (47).

The main purpose of this scoping review was to ascertain whether the hormones of the thyroid axis vary during weight loss in overweight/obese children/adolescents. The examination of the 16 selected studies allowed us to conclude that most of the results pointed to the absence of elevated levels at baseline, in agreement with a previous review (5). Also, most of the studies showed that the changes in body composition parameters in response to the multidisciplinary interventions correlated positively with fT3/TT3/TSH. Further studies are necessary to elucidate the relevance of the variations in hormone levels to the establishment of overweight/obesity and to the recovery from these conditions in children/adolescents. With respect to fT4/TT4, the most common finding was of unchanged levels and hence, no significant association with weight loss. Importantly, the response to the intervention has even been found to not be affected by fT4 supplementation.

## Data Availability

N/A

## Author Contributions

C.R.R. and E.B.R.; methodology, C.R.U., E.B.R., A.C.F.C. and M.C.P.F; review per peers, C.R.U., A.P.P., F.T., and E.B.R.; formal analysis, C.R.U., A.P.P. F.T., M.M.T., A.L.S., and E.B.R.; writing—original draft preparation, C.R.U., E.B.R. and M.C.P.F.; writing—review and editing, C.R.U., E.B.R and M.M.T.. All authors have read and agreed to the published version of the manuscript.”

## References

1. World Health Organization (WHO). Guideline: assessing and managing children at primary health-care facilities to prevent overweight and obesity in the context of the double burden of malnutrition. Updates for the Integrated Management of Childhood Illness (IMCI). [Internet]. 2017 [cited 2019 Jul 18]. Available from: http://apps.who.int/bookorders.

2. NCD Risk Factor Collaboration (NCD-RisC). Worldwide trends in body-mass index, underweight, overweight, and obesity from 1975 to 2016: a pooled analysis of 2416 population-based measurement studies in 128·9 million children, adolescents, and adults. Lancet [Internet]. 2017 [cited 2019 Mar 11];390:2627–42. Available from: http://dx.doi.org/10.1016/

3. Abdullah A, Aceh B, Courten M De, Stevenson CE, Walls H. The number of years lived with obesity and the risk of all-cause and cause-specific mortality. Int J Epidemiol. 2011;50(2):985–96.

4. Mullur R, Liu YY, Brent GA. Thyroid hormone regulation of metabolism. Physiol Rev. 2014;94(2):355–82.

5. Witkowska-Sedek E, Malgorzata Ruminska AK. Thyroid dysfunction in obese and overweight children. Endokrynol Pol. 2017;68(1):54–60.

6. Hoelscher DM, Kirk S, Ritchie L, Cunningham-Sabo L. Position of the Academy of Nutrition and Dietetics: Interventions for the Prevention and Treatment of Pediatric Overweight and Obesity. J Acad Nutr Diet. 2013;113:1375–94.

7. Mcgowan J, Sampson M, Salzwedel DM, Cogo E, Foerster V, Lefebvre C. GUIDELINE STATEMENT PRESS Peer Review of Electronic Search Strategies : 2015 Guideline Statement. J Clin Epidemiol. 2016;75:40–6.

8. Kiortsis DN, Durack I, Turpin G. Effects of a low-calorie diet on resting metabolic rate and serum tri-iodothyronine levels in obese children. Eur J Pediatr. 1999;158(6):446–50.

9. Rijks JM, Plat J, Dorenbos E, Penders B, Gerver WJM, Vreugdenhil ACE. Association of TSH with cardiovascular disease risk in overweight and obese children during lifestyle intervention. J Clin Endocrinol Metab. 2017;102(6):2051–8.

10. Aeberli I, Jung A, Murer SB, Wildhaber J, Wildhaber-Brooks J, Knöpfli BH, et al. During Rapid Weight Loss in Obese Children, Reductions in TSH Predict Improvements in Insulin Sensitivity Independent of Changes in Body Weight or Fat. J Clin Endocrinol Metab. 2010;95(12):5412–8.

11. Shalitin S, Yackobovitch-Gavan M, Phillip M. Prevalence of thyroid dysfunction in obese children and adolescents before and after weight reduction and its relation to other metabolic parameters. Horm Res. 2009;71(3):155–61.

12. Baş VN, Aycan Z, Ağladioğlu SY, Kendirci HNP. Prevalence of hyperthyrotropinemia in obese children before and after weight loss. Eat Weight Disord. 2013;18(1):87–90.

13. Wolters B, Lass N, Reinehr T. TSH and free triiodothyronine concentrations are associated with weight loss in a lifestyle intervention and weight regain afterwards in obese children. Eur J Endocrinol. 2013;168(3):323–9.

14. Licenziati MR, Valerio G, Vetrani I, Maria G De, Liotta F, Radetti G. Altered Thyroid Function and Structure in Children and Adolescents Who Are Overweight and Obese: Reversal After Weight Loss. J Clin Endocrinol Metab. 2019;104(7):2757–65.

15. Bouglé D, Morello R, Brouard J. Thyroid function and metabolic risk factors in obese youth. Changes during follow-up: a preventive mechanism? Exp Clin Endocrinol diabetes. 2014;122(9):548–52.

16. Radetti G, Longhi S, Baiocchi M, Cassar W, Buzi F. Changes in lifestyle improve body composition, thyroid function, and structure in obese children. J Endocrinol Invest. 2012;35(3):281–5.

17. Marras V, Casini MR, Pilia S, Carta D, Civolani P, Porcu M, et al. Thyroid function in obese children and adolescents. Horm Res Paediatr [Internet]. 2010;73(3):193–7. Available from: https://www.scopus.com/inward/record.uri?eid=2-s2.0-77951276542&doi=10.1159%2F000284361&partnerID=40&md5=12905e0603e2ec9b14938e68ac15537d

18. Cayir A, Doneray H, Kurt N, Orbak Z, Kaya A, Turan MI, et al. Thyroid functions and trace elements in pediatric patients with exogenous obesity. Biol Trace Elem Res. 2014;157(2):95–100.

19. Reinehr T, Sousa G De, Andler W. Hyperthyrotropinemia in Obese Children Is Reversible after Weight Loss and Is Not Related to Lipids. J Clin Endocrinol Metab. 2006;91(8):3088–91.

20. Reinehr T, Andler W. Thyroid hormones before and after weight loss in obesity. Arch Dis Child. 2002;87(4):320–3.

21. Butte NF, Brandt ML, Wong WW, Liu Y, Mehta NR, Wilson TA, et al. Energetic adaptations persist after bariatric surgery in severely obese adolescents. Obesity. 2015;23(3):591–601.

22. Eliakim A, Barzilai M, Wolach B, Nemet D. Should we treat elevated thyroid stimulating hormone levels in obese children and adolescents? Int J Pediatr Obes. 2006;1(4):217–21.

23. Kumar S, Dayal D, Attri SV, Gupta A, Bhalla AK. Levothyroxine supplementation for obesity-associated thyroid dysfunction in children: A prospective, randomized, case control study. Pediatr Endocrinol Diabetes Metab. 2019;25(3):107–13.

24. Soydan L, Ozturk E, Onal Z C N., Associations of Thyroid Volume and Function with Childhood Obesity. Acta Endocrinol. 2019;15(1):123–8.

25. Stichel H, L’allemand D, Grüters A. Thyroid function and obesity in children and adolescents. Horm Res. 2000;54(1):14–9.

26. Lobotková D, Staníková D, Staník J, Červeňová O, Bzdúch V, Tichá L. Lack of association between peripheral activity of thyroid hormones and elevated TSH levels in childhood obesity. J Clin Res Pediatr Endocrinol. 2014;6(2):100–4.

27. Ayala-Moreno M del R, Guerrero-Hernández J, Vergara-Castañeda A, Salazar-Aceves G, Cruz-Mercado DE. Thyroid function in pediatric population with different nutritional status. Bol Med Hosp Infant Mex. 2018;75(5):279–86.

28. Ghergherehchi R, Hazhir N. Thyroid hormonal status among children with obesity. Ther Adv Endocrinol Metab. 2015;6(2):51–5.

29. Wang Y, Dong X, Fu C, Su M, Jiang F, Xu D. Thyroid Stimulating Hormone (TSH) Is Associated With General and Abdominal Obesity: A Cohort Study in School-Aged Girls During Puberty in East China. Front Endocrinol (Lausanne). 2020;11(9):1–9.

30. Zhang L, Wang G. Association Between Body Mass Index and Thyroid Function in Euthyroid Chinese Adults. Med Sci Monit. 2021;27(27):1–8.

31. Mwafy S, Yassin M, Mousa R. Thyroid hormones, lipid profile and anthropometric changes after programmed weight loss in Palestinian obese adult females. Diabetes Metab Syndr Clin Res Rev. 2018;12(3):269–73.

32. Sari R, Balci MK, Altunbas H, Karayalcin U. The effect of body weight and weight loss on thyroid volume and function in obese women. Clin Endocrinol (Oxf). 2003;59(2):258–62.

33. Dursun F, Atasoy Öztürk T, Seymen Karabulut G, Klrmlzlbekmez H. Obesity-related thyroiditis in childhood: Relationship with insulin resistance. J Pediatr Endocrinol Metab. 2019;32(5):471–8.

34. Joshi SR. Thyro-weight: Unlocking the link between thyroid disorders and weight. J Assoc Physicians India. 2018;66(March):70–3.

35. Liu G, Liang L, Bray GA, Qi L, Hu FB, Rood J, et al. Thyroid hormones and changes in body weight and metabolic parameters in response to weight loss diets: The POUNDS LOST trial. Int J Obes. 2017;41(6):878–86.

36. Chen H, Zhang H, Tang W, Xi Q, Liu X, Duan Y, et al. Thyroid function and morphology in overweight and obese children and adolescents in a Chinese population. J Pediatr Endocr Met 2013; 2013;26(5–6):489–96.

37. Roef GL, Rietzschel ER, Van Daele C, Taes YE, De Buyzere M, Gillebert TC, et al. Triiodothyronine and free thyroxine levels are differentially associated with metabolic profile and adiposity-related cardiovascular risk markers in euthyroid middle-aged subjects. Thyroid. 2014;24(2):1–31.

38. Natah TM, Wtwt MA, Hadi MA, Farhood HF. Thyroid Metabolic Hormones and its Correlation with BMI and Lipid Profile in Healthy People. Food Sci Qual Manag. 2013;18:18–25.

39. Fontenelle LC, Feitosa MM, Severo JS, Freitas TEC, Morais JBS, Torres-Leal FL, et al. Thyroid Function in Human Obesity: Underlying Mechanisms. Horm Metab Res. 2016;48(12):787–94.

40. Bastemir M, Akin F, Alkis E, Kaptanoglu B. Obesity is associated with increased serum TSH level, independent of thyroid function. Swiss Med Wkly [Internet]. 2007 [cited 2018 Sep 14];137(29–30):431–4. Available from: https://smw.ch/en/resource/jf/journal/file/view/article/smw/en/smw.2007.11774/smw.2007.11774.pdf/

41. Hoermann R, Midgley JEM, Larisch R, Dietrich JW. Homeostatic Control of the Thyroid–Pituitary Axis: Perspectives for Diagnosis and Treatment. Front Endocrinol (Lausanne) [Internet]. 2015 Nov 20 [cited 2018 Nov 26];6:177. Available from: http://journal.frontiersin.org/Article/10.3389/fendo.2015.00177/abstract

42. Fox CS, Pencina MJ, D’Agostino RB, Murabito JM, Seely EW, Pearce EN, et al. Relations of Thyroid Function to Body Weight. Arch Intern Med [Internet]. 2008;168(6):587–92. Available from: https://jamanetwork-com.rsm.idm.oclc.org/journals/jamainternalmedicine/fullarticle/414105

43. Agnihothri R V, Courville AB, Linderman JD, Smith S, Brychta R, Remaley A, et al. Moderate Weight Loss Is Sufficient. Thyroid. 2014;24(1).

44. Marzullo P, Minocci A, Mele C, Fessehatsion R, Tagliaferri M, Pagano L, et al. The relationship between resting energy expenditure and thyroid hormones in response to short-term weight loss in severe obesity. PLoS One [Internet]. 2018 [cited 2018 Nov 26];13(10):1–12. Available from: https://doi.org/10.1371/journal.pone.0205293

45. Rosenbaum M, Hirsch J, Murphy E, Leibel RL. Effects of changes in body weight on carbohydrate metabolism, catecholamine excretion, and thyroid function 1 – 4. Am J Clin Nutr. 2018;71(March):1421–32.

46. Muller M, Enderle J, Pourhassan M, Braun W, Eggeling B, Lagerpusch M. Metabolic adaptation to caloric restriction and subsequent refeeding : the Minnesota Starvation Experiment revisited 1, 2. Am J Clin Nutr. 2015;102:1–13.

47. Johnstone AM, Murison SD, Duncan JS, Rance KA, Speakman JR. Factors influencing variation in basal metabolic rate include fat-free mass, fat mass, age, and circulating thyroxine but not sex,. Am J Clin Nutr. 2005;82(March):941–8.

